# The role of M1 to M2 macrophage polarization in the etiology of idiopathic gastroparesis: GWAS perspective

**DOI:** 10.1101/2022.06.22.22276605

**Authors:** Sandra P. Smieszek, Jesse L. Carlin, Changfu Xiao, Christos M. Polymeropoulos, Gunther Birznieks, Mihael H. Polymeropoulos

## Abstract

Gastroparesis is a serious medical condition characterized by delayed gastric emptying and symptoms of nausea, vomiting, bloating, fullness after meals, and abdominal pain. An innate immune dysregulation and injury to the interstitial cells of Cajal and other components of the enteric nervous system are likely central to the pathogenesis of gastroparesis. Thus far, little is known about the underlying genetic risk factors for gastroparesis. To ascertain these genetic risk factors for gastroparesis we conducted the first large whole-genome sequencing genome-wide association study (GWAS) analysis of gastroparesis patients.

The GWAS focused on idiopathic and diabetic gastroparesis cases as compared to matched controls as well as compared idiopathic versus diabetic cases. Amongst the top variants, we report a novel genetic risk variant for idiopathic gastroparesis - genetic association of the *Solute Carrier Family 15 (SLC15)* locus. This signal is driven by multiple variants with top missense variant SLC15A4:NM145648:exon2:c.T716C:p.V239A, rs33990080. SLC15A4 was shown to mediate M1-prone metabolic shifts in macrophages and guards immune cells from metabolic stress. The risk variant carriers have a significantly higher nausea score at baseline. We also delineated a number of statistically significant loci including novel ones as well as previously known loci such as *HLA-DQB1* - rs9273363, variant associated with Type 1 diabetes..

The GWAS picture that is emerging implicates the role of the machinery of M1 to M2 macrophage polarization in the etiology of idiopathic gastroparesis. We suggest that macrophage polarization fate could lead to the destruction of the interstitial cells of Cajal which effectively leads to abnormal gastric emptying.

## Introduction

Gastroparesis is a serious medical condition characterized by delayed gastric emptying in the absence of mechanical obstruction^1^. The main symptoms include nausea, vomiting, bloating, fullness after meals, and abdominal pain^1^. The overall standardized prevalence of gastroparesis is 267.7 per 100,000 US adults, whereas the prevalence of “definite” gastroparesis is 21.5 per 100,000 ^2^. Common etiologies include diabetes, post-surgical gastroparesis and idiopathic gastroparesis (with estimates of idiopathic (∼11.3%) vs diabetic (>51.7%))^2^.

An innate immune dysregulation and injury to the interstitial cells of Cajal (ICC) and other components of the enteric nervous system through paracrine and oxidative stress mediators are likely central to the pathogenesis of gastroparesis^3^. Loss of antral CD206 positive anti-inflammatory macrophages is a key feature in human gastroparesis and it associates with ICC loss^4^. A review of the literature suggests that oxidative stress associated with diabetes activates macrophages. Activation of CD206 +, anti-inflammatory M2 macrophages expressing heme oxygenase 1 (HO1) is protective while activation of pro-inflammatory M1 macrophages which lack HO1 is injurious and leads to the development of delay in gastric emptying’^5^. Low numbers of M2 macrophages were correlated with loss of ICC and were associated with both idiopathic and diabetic gastroparesis^4^. Importantly, when the anti-inflammatory M2 macrophages were switched to pro-inflammatory M1 macrophages, delayed GE was evident in animal models^4^.

Consistent with this notion is gastroparesis in patients receiving immune checkpoint inhibitors – blockage to PD-1/PD1-L and the resultant damage to the anti-inflammatory macrophages^6^. It is plausible that the blockage of PD-1/PD-L1 by ICI contributes to the onset of gastroparesis as a result of an immune-related adverse event. This is consistent with the decreased production of protective cytokines by M2 macrophages which was shown in acute lung injury with the blockage of PD-1/PD1-L^6^.

Thus far, there little is known regarding the common genetic risk factors for idiopathic gastroparesis. Prior studies reported Longer poly-GT repeats in the HMOX1 gene are more common in African Americans with gastroparesis, particularly in patients with diabetes^7^. To ascertain the genetic risk factors for gastroparesis we conducted a large whole-genome sequencing (WGS) of samples obtained from well clinically characterized gastroparesis studies of both diabetic and idiopathic gastroparesis. We have done a GWAS comparing idiopathic and diabetic cases with controls as well l as a GWAS on idiopathic versus with diabetic cases. We have delineated the expected loci in the diabetic gastroparesis cohort and furthermore, we report novel associations with the idiopathic gastroparesis.

## Results

Whole-genome sequencing was done on 1385 samples obtained from consented gastroparesis patients. Samples were obtained from patients participating in two gastroparesis studies phase II clinical study (VP-VLY-686-2301 and ongoing phase III gastroparesis clinical study (VP-VLY-686-3301). Inclusion criteria included patients with idiopathic or diabetic gastroparesis with moderate to severe nausea, delayed gastric emptying, daily 50% worst average nausea score ≥3 and GCSI nausea score ≥ 3 at screening. In the phase III study subjects included male and female adults ages 18-70 with a diagnosis of diabetic or idiopathic gastroparesis. Subjects enrolled in the study had evidence of delayed gastric emptying, and moderate to severe nausea, daily average nausea score of > 2.5, at least one episode of vomiting, PAGI-SYM nausea score of ≥ 2 at screening, and controlled blood glucose levels with HbA1c < 10%. Demographics and clinical characteristics are provided in **Table 1**. Self-reported ancestry was further confirmed with Principal Component Analysis (PCA) - details provided in the Methods section. Association results were obtained from logistic models for the 4 following GWAS analyses: idiopathic vs. controls, diabetic vs. controls, gastroparesis cases vs. non gastroparesis matched controls and diabetic vs. idiopathic cases.

### Idiopathic Gastroparesis

To avoid spurious signals resulting from population substructure, the largest set of EUR ancestry was used for the purpose of the idiopathic and diabetic analyses. The analysis consisted of variants with Minor Allele Frequency (MAF) >1, EUR individuals (PCA-defined) in both cases (n=319) and controls (n=896). For comparisons of variant frequencies, we used both gastroparesis cohorts, controls and gnomAD^8^ (matched by ancestry as defined by PCA). Cases and controls were called by uniform GATK pipeline, with only variants passing GATK’s Variant Quality Score Recalibration – (VQSR) and internal MAF cutoffs were analyzed. Amongst the top strongest loci is a signal from variant within the Solute carrier family 15 (SLC15A4) gene - lead variant rs10847696 (*p*-value<0.000004 (OR:1.9). The non-synonymous variant of interest, NM145648:exon2:c.T716C:p.V239A, rs33990080 is also depicted on the Manhattan plot **Figure 1A** and the SLC15A4 regional zoom plot **Figure 2**. We report 69 carriers among 214 idiopathic patients versus 165 out of 896 ancestry-matched controls (MAF: cases 0.18; MAF: controls 0.09) and OR: 1.9. The MAF of the top coding variant among all gastroparesis patients, combining both diabetic and idiopathic patients is 0.16. The global MAF among non-finnish-europeans (NFE) in a public database (gnomAD^8^) is 0.09. The significant effect persists when idiopathic cases are compared with diabetic cases as the MAF of diabetic cases (0.10) is comparable to that of population controls (0.09). The variant is a highly statistically significant (*p*-value< 10^^-17^) eQTL for chr12 *GLT1D1* and *SLC15A4* (GTEX^9^) **Supplementary Figure 1A**. The identified variant carriers have a significantly higher nausea score at baseline with a mean difference ∼0.3 unit, and ∼0.6 unit if comparing homozygotes (*p*-value<0.04) as shown on **Supplementary Figure 1B**. There are additional significant variants reported within *SLC15A4* gene in association with idiopathic gastroparesis further supporting the effect carried by identified region. The top results for the idiopathic vs. controls analysis are presented in **Table 2** as well as in the **Supplementary Material Data**. Other identified loci include intergenic variants in the region of *MIR8054* and *LUZP2* genes as well as other loci depicted in Table 2. Importantly, we have furthermore replicated the effect of this *SLC15A4* variant in another batch of EUR gastroparesis samples with OR 1.6 *p*-value<10^5.

**Table 2.**
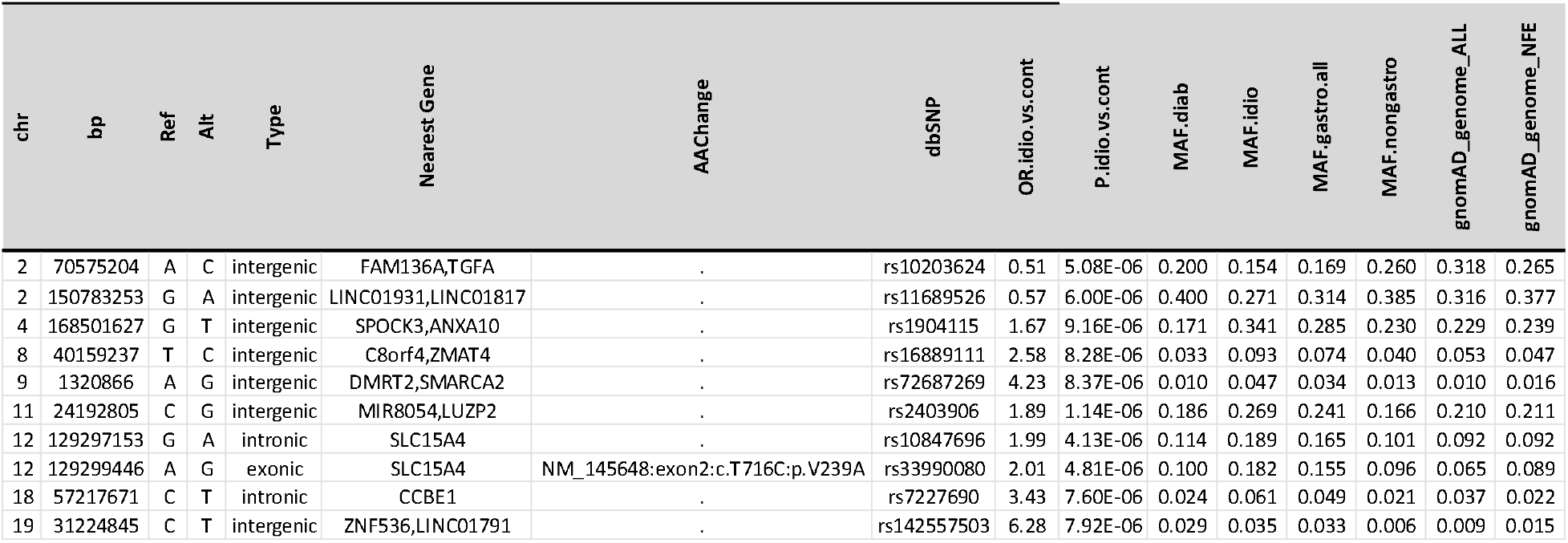
Top GWAS variants for idiopathic cases versus controls

**Figure 1.**
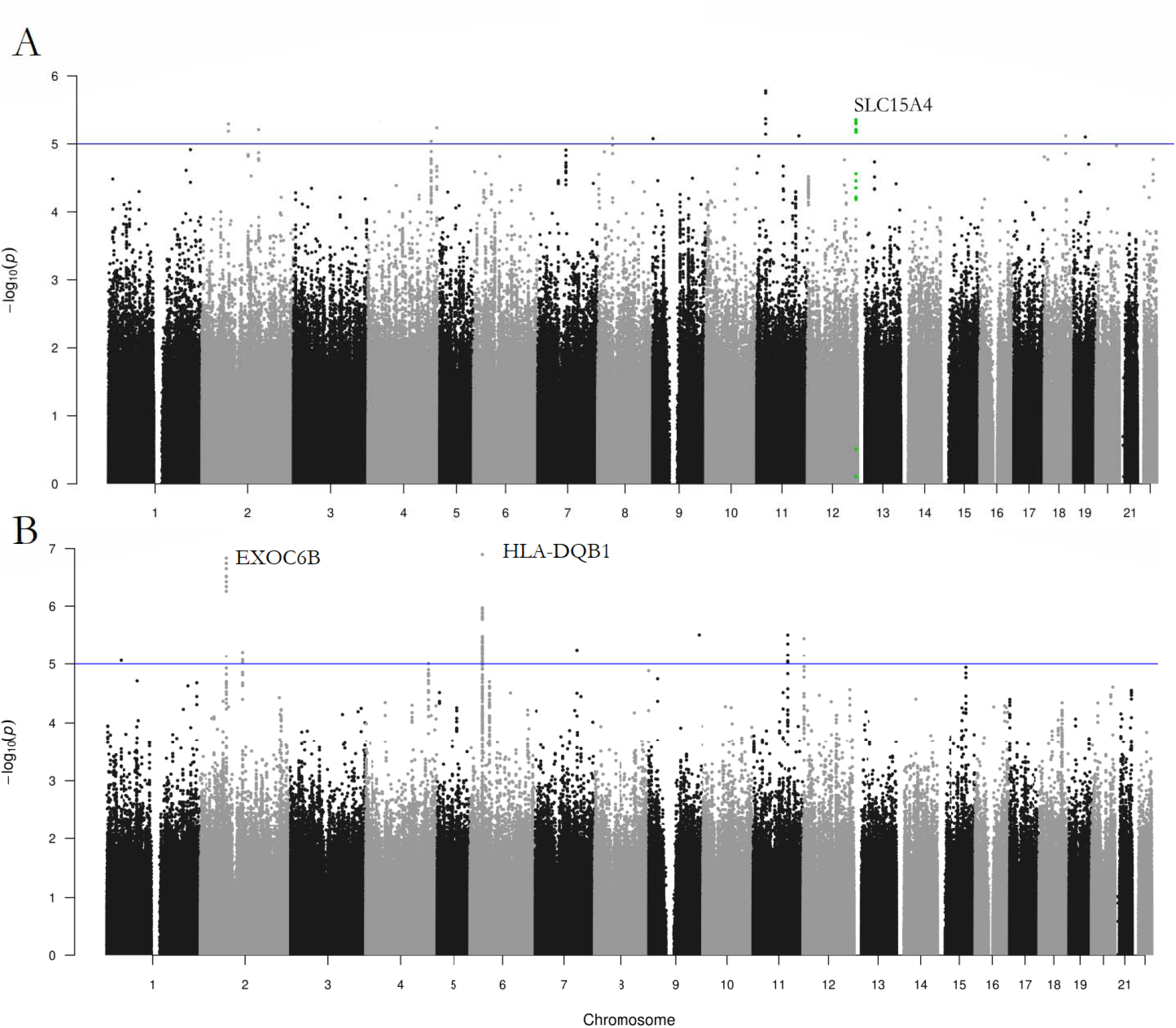
Manhattan plots showing top results for A. GWAS: idiopathic gastroparesis versus controls B. GWAS: diabetic gastroparesis cases versus controls

**Figure 2.**
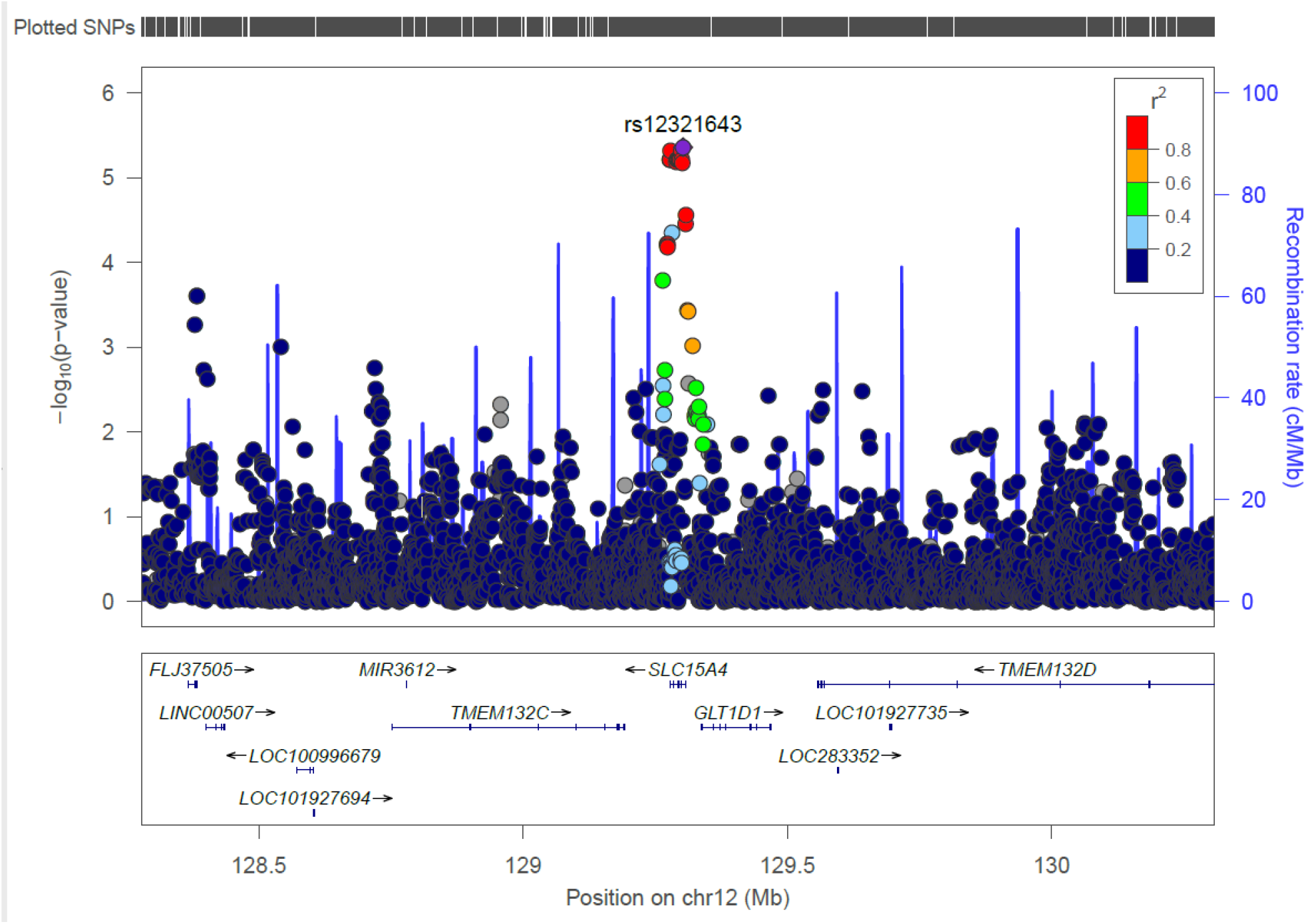
Zoom plot showing *SLC15A4* rs33990080 variant of region detected in the idiopathic gastroparesis cases versus matched controls analysis

### Diabetic vs. Idiopathic Gastroparesis

Whereas the primary focus of this analysis was on idiopathic rather than diabetic gastroparesis, both sets of analysis yielded informative results. In this set idiopathic controls are super controls as this is a set with a confirmed negative diabetes diagnosis. Top variants in diabetic gastroparesis GWAS included variants pointing to *HLA-DQB1* with lead variant rs9275638. Among the reported top variants is rs9273363 - a known risk factor for Type 1 diabetes, with an OR of 2.1, *p*-value<10^^-5^, a common variant with a MAF of 0.39 in diabetic gastroparesis cases versus MAF of 0.22 in idiopathic controls as well as MAF 0.27 in matched EUR controls. The variant is tagging the HLA DQB1*03:02 haplotype and the 6q22.33 region, which contains the genes encoding protein tyrosine phosphatase receptor k (*PTPRK*). Amongst the other top variants are loci within *EXOC6B* depicted in **Figure 1B** and **Table 3**. *EXOC6B*’s top variant is a UTR variant rs41416 highly increased among diabetic but not idiopathic cases (OR 3.3, *p-value<10*^*^-7*^). This exocyst complex gene has been previously associated with increased risk of Type II diabetes. We have furthermore calculated a T2D polygenetic risk score to see how well we can cluster cases. Polygenic scores were constructed for diabetes by summing the number of risk alleles carried by each individual, weighted by the effect size estimates from well-established genome-wide significant associations derived from Li et al., 2021 (60 SNPs)^10^. We differentiate idiopathic from diabetic gastroparesis with a diabetes genetic risk score, - statistically significant result (*p*-value<0.0004). This reinforces the capacity to subgroup cases based on diabetes risk score. This is presented in the **Supplementary Material Figure 2** Additionally, we explored the frequencies of previously reported variants in a functional dyspepsia GWAS^11^. Two variants previously reported came out as nominally significant: rs2595968 (NFASC *p*-value<0.004) and rs17597505 (*p*-value<0.03).

**Table 3.** Top GWAS variants for diabetic cases versus idiopathic cases

| chr | bp | Ref | Alt | Type | Nearest Gene | ExonicFunc | dbSNP | OR.logit.diab.vs.idio | P.logit.diab.vs.idio | MAF.diab | MAF.idio | MAF.gastro.all | MAF.nongastro | gnomAD_genome_ALL | gnomAD_genome_NFE |
| --- | --- | --- | --- | --- | --- | --- | --- | --- | --- | --- | --- | --- | --- | --- | --- |
| 2 | 72416952 | TCTC | - | intronic | EXOC6B | . | rs139115191 | 3.36 | 1.45E-07 | 0.291 | 0.122 | 0.177 | 0.184 | 0.115 | 0.164 |
| 2 | 114998082 | G | C | intergenic | LINC01191,DPP10 | . | rs3132039 | 3.34 | 6.09E-06 | 0.229 | 0.096 | 0.140 | 0.120 | 0.090 | 0.129 |
| 6 | 32698988 | A | C | intergenic | HLA-DQB1,HLA-DQA2 | . | rs7751856 | 3.03 | 1.26E-07 | 0.610 | 0.435 | 0.492 | 0.462 | 0.489 | 0.540 |
| 6 | 32729642 | C | G | exonic | HLA-DQB2 | synonymous SNV | rs762815 | 2.5 | 3.45E-06 | 0.55 | 0.4 | 0.45 | 0.41 | 0.66 | 0.57 |
| 11 | 95405933 | C | T | intergenic | LOC100129203,FAM76B | . | rs7931781 | 2.40 | 3.08E-06 | 0.595 | 0.409 | 0.470 | 0.479 | 0.561 | 0.538 |
| 12 | 2964735 | G | A | ncRNA_intronic | LOC100507424 | . | rs7138715 | 2.86 | 3.55E-06 | 0.286 | 0.143 | 0.190 | 0.191 | 0.263 | 0.176 |
| 15 | 78567853 | G | T | exonic | DNAJA4 | synonymous SNV | rs967821 | 2.79 | 1.15E-05 | 0.286 | 0.150 | 0.194 | 0.175 | 0.212 | 0.193 |

## Discussion

Whole-genome sequencing can provide unique insights into genetic determinants of gastroparesis, by uncovering associations between common as well as rare genetic variants. Such loci can be extremely helpful, because they are often coding, thereby pointing directly to a causal gene. In this study focused primarily on idiopathic and diabetic gastroparesis cases. We report novel variants associated with idiopathic gastroparesis as well as known risk factors previously associated with diabetes. Importantly, we report a novel genetic association of the *SLC15A4* (p.Val239Ala) risk variant conferring a greater risk of idiopathic gastroparesis.

Solute carrier family 15 (SLC15) A4 is a lysosome-resident, amino acid/oligopeptide transporter that is preferentially expressed in immune cells^12^. This proton-coupled amino-acid transporter mediates the transmembrane transport of L-histidine and some di- and tripeptides from inside the lysosome to the cytosol, and plays a key role in innate immune responses. Importantly, it is required for TLR7/9-dependent type I interferon production^13^. SLC15A4 has been associated with disorders such as inflammatory bowel diseases and systemic lupus erythematosus^13^,^14^. High levels of *SLC15A4* transcripts were observed in human antigen-presenting cells, including dendritic cells, activated macrophages, and B cells^13^. SLC15A4 was shown to mediate M1-prone metabolic shifts in macrophages and guards immune cells from metabolic stress^12^. SLC15A4 loss disturbed the coupling of glycolysis and the TCA cycle, and SLC15A4-deficient macrophages preferred to use glutamine rather than glucose as a carbon source for the TCA cycle^12^. SLC15A4-deficient macrophages produced low levels of itaconate and proinflammatory IL-12 cytokine members. Correspondingly, *SLC15A4*^−/−^ mice developed a less severe form of Th1-dependent colitis than *SLC15A4*^+/+^ mice^13^. SLC15A4 was shown to promote colitis through Toll-like receptor 9 and NOD1-dependent innate immune responses. It is involved in maintaining the histidine homeostasis within intracellular compartments and is crucial for eliciting effective innate immune responses. SLC15A4-intact but not SLC15A4-deficient macrophages became resistant to fluctuations in environmental nutrient levels by limiting the use of the glutamine source; thus, SLC15A4 was critical for macrophage’s respiratory homeostasis^12^.

Previous reports have shown that when the anti-inflammatory M2 macrophages are switched to pro-inflammatory M1 macrophages, delayed GE was evident in animal models ^4^. This is consistent with the GWAS picture that is emerging where the machinery of M1 to M2 macrophage polarization seems to be implicated. These findings suggest a mechanism of metabolic regulation in which an amino acid transporter acts as a gatekeeper that protects immune cells’ ability to acquire an M1-prone metabolic phenotype in inflammatory tissues by mitigating metabolic stress. It seems the detected loci could be directly connected to macrophage polarization either via gain of function of hyperexpression, mechanisms that still need to be confirmed. Past reports on idiopathic cases suggested a reduction in both myenteric and intramuscular interstitial cells of Cajal at the center of the etiology of gastroparesis^15^. Successful gastric emptying results from the successful cooperation between smooth muscle, enteric and autonomic nerves, and ICC^15^. It may be the case that multiple paths lead to the same aberrant pathway such as in the M1/M2 macrophage polarization leading to gastroparesis, both of the diabetic or idiopathic etiology. In this report, we show such potential risk factors from a genetic perspective. We suggest that macrophage polarization fate could lead to the destruction of the interstitial cells of Cajal effectively leads to abnormal gastric emptying.

Nevertheless, our findings despite initial replication require further confirmation as the number of sequenced cases increases. Large association studies require careful control for population stratification and hence analysis was focused on EUR samples. Further analyses are necessary across other ancestries. There are other variants delineated in this study that call for further confirmation and research into their mechanisms of action. By depleting the diabetic cases and doing a sub-analysis we created a homogenous set for exploratory GWAS. We successfully detected risk for HLA-DQB1 – in the diabetes cohort - we also identified other loci that call for further confirmatory studies such as EXOC6B. Despite this being the largest WGS study reported, size is still a limitation and further replication of these findings will be necessary.

In summary, we detected novel loci associated with idiopathic gastroparesis. We also reproduced a genome-wide significant signal for diabetic gastroparesis – a whole-genome-wide significant association between HLA-DQB1 and Type 1 diabetic gastroparesis amongst other variants. Our results also suggest an association between macrophage polarization at the center of the etiology of idiopathic gastroparesis, a mechanism that has also been implicated in the etiology of gastroparesis in patients with diabetes. Future genome-wide studies of rare variants will require a larger sample size to fully understand the genetic architecture of gastroparesis.,

## Methods

### Datasets

Samples were obtained from patients participating in two respective gastroparesis clinical trials phase II clinical study – VP-VLY-686-2301^16^ and phase III clinical study VP-VLY-686-3301. Inclusion criteria for both studies included patients with idiopathic or diabetic gastroparesis with moderate to severe nausea and delayed gastric emptying, as well as controlled diabetes status. The first gastroparesis dataset (phase II clinical study – VP-VLY-686-2301) was a multiethnic cohort (n -119)^16^. This cohort was composed of 90.7% females; the average age was 45.9; 86.2% of the patients self-identified as White, 10.5% self-reported as Black or African American, 2% self-reported American Indian or Alaska Native and 1.3% reported as Asian. The second study (VP-VLY-686-3301) was a multiethnic cohort composed of 70.2% females; the average age was 48.5; 64.5% of the patients self-identified as White, 31.7% self-reported as Black or African American, 0.8% self-reported American Indian or Alaska Native and 1.9% reported as Asian.

### Genetic analysis

DNA samples were quantified using fluorescent-based assays (PicoGreen) to accurately determine whether sufficient material is available for library preparation and sequencing. DNA sample size distributions were profiled by a Fragment Analyzer (Advanced Analytics) or BioAnalyzer (Agilent Technologies), to assess sample quality and integrity. WGS libraries were prepared using the Truseq DNA PCR-free Library Preparation Kit. Whole Genome data were processed on NYGC automated pipeline. Paired-end 150 bp reads were aligned to the GRCh37 human reference (BWA-MEM v0.7.8) and processed with GATK best-practices workflow (GATK v3.4.0). The mean coverage was 35.8reflecting the average of the samples. All high-quality variants obtained from GATK pipeline were functionally annotated (intronic, intergenic, splicing, nonsynonymous, stopgain and frameshifts) based on RefSeq transcripts. Cases and controls were called with the sample pipeline, only variants passing GATK’s Variant Quality Score Recalibration - VQSR) internal MAF cutoff were included in the analysis.

### GWAS

We performed a GWAS using variants with minor allele frequencies higher than 1% and allele count of 10 or more, with the same analysis design used in the landmark COVID-19 HGI GWAS^17^. We performed single variant association tests using a GWAS additive model framework. Logistic models adjusted for PC, age and sex were conducted in PLINK^18^. The analysis was limited to individuals of European (EUR) genetic ancestry due to the largest cohort available in this ancestry and in order to avoid confounders in the form of population substructure. The Principal Component Analysis and PC definitions are based on the one provided by the COVID-19 Host Genetics Initiative, available here for reference: https://github.com/covid19-hg/pca_projection. A common set of SNPs was used for ancestry definitions to match those for 1000 genomes project (V3).

## Data Availability

upon request

---

Table 1. Demographics and clinical characteristics of the screened cohort of gastroparesis patients

## Notes

### Competing Interest Statement

The authors are employees of Vanda Pharmaceuticals Inc.

### Funding Statement

This study was funded by Vanda Pharmaceuticals Inc.

### Author Declarations

Advarra IRB Pro00032689

