## Supplementary Material for "The role of M1 to M2 macrophage polarization in the etiology of idiopathic gastroparesis: GWAS perspective"

Smieszek et al., 2022

Figure 1A and 1B

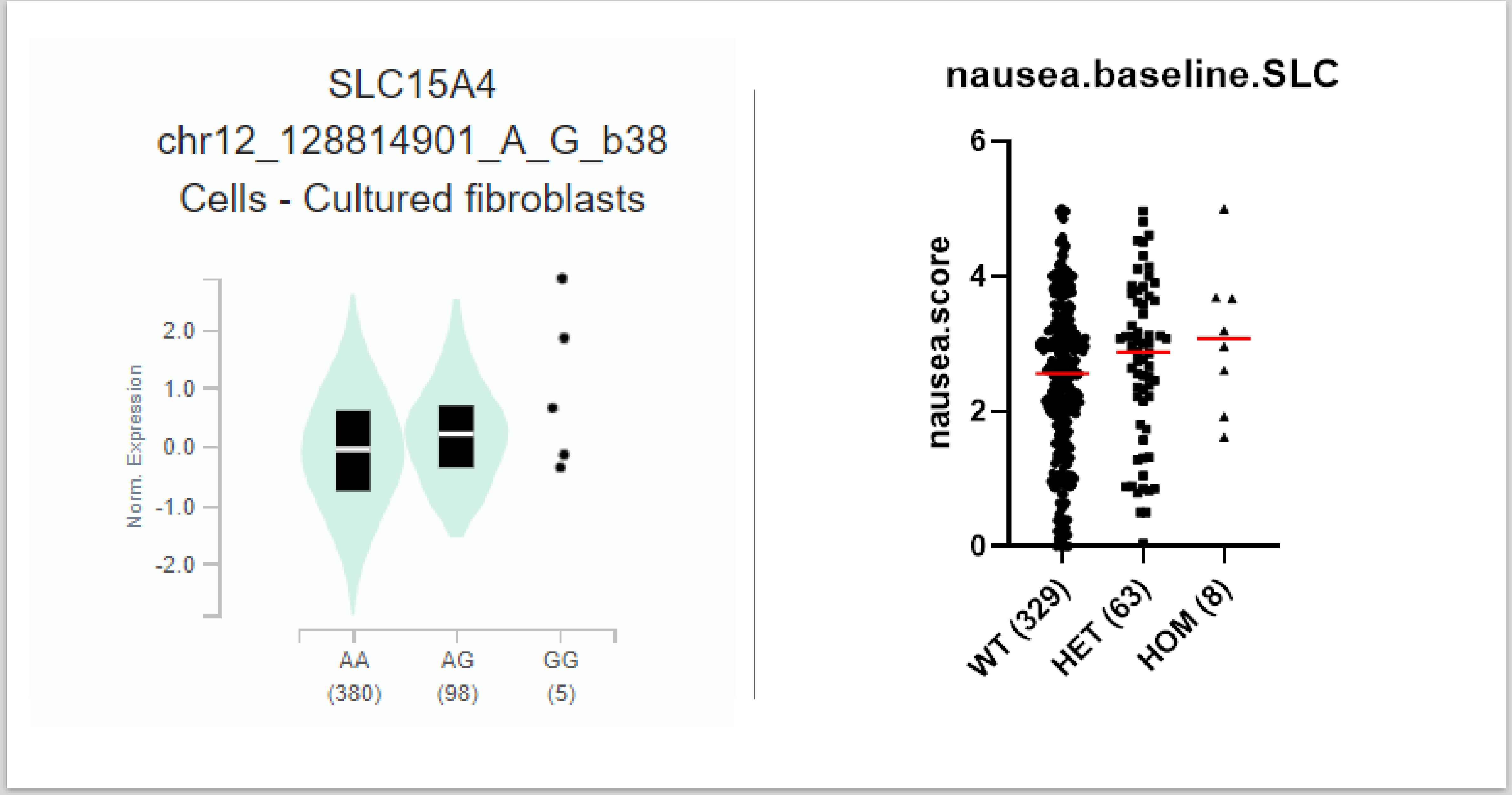

### Figure 2A and 2B: Genetic risk score

- Polygenic scores were constructed for diabetes by summing the number of risk alleles carried by each individual, weighted by the effect size estimates from well-established genome-wide significant associations derived from Li et al., (60 SNPs)
- We differentiate idiopathic from diabetic gastroparesis with a diabetes genetic risk score, - result that is statistically significant (p-value<0.0004).
- We also recapitulate a number of known significant loci implicated in diabetes risk including (HLA-DQB1,HLA-DQA2, EXOC6B) in the idiopathic versus diabetic gastroparesis analysis.

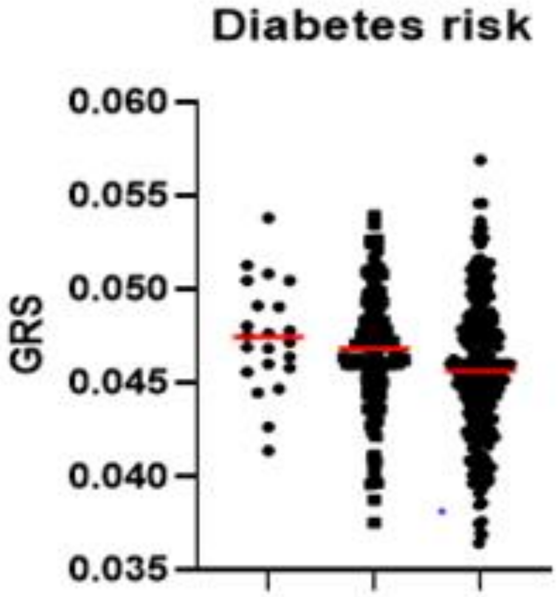


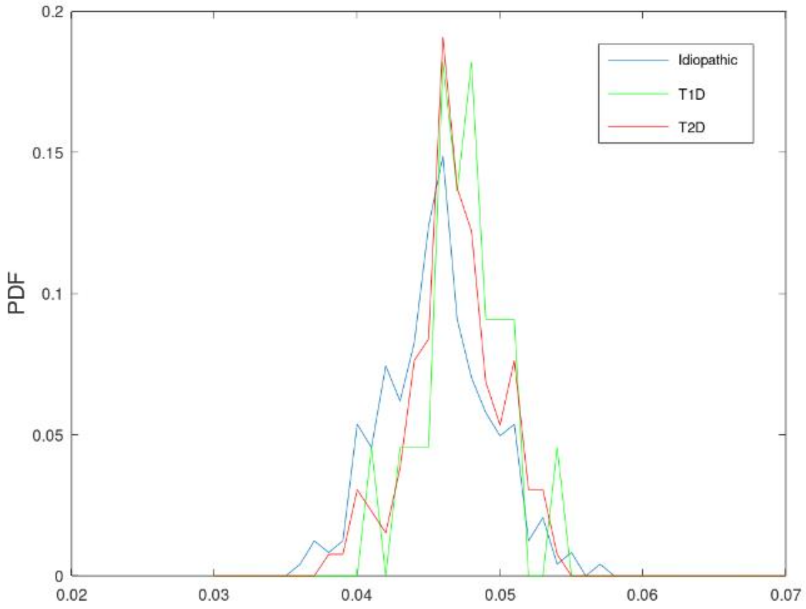

### SLC15A4 ns variant **rs33990080**

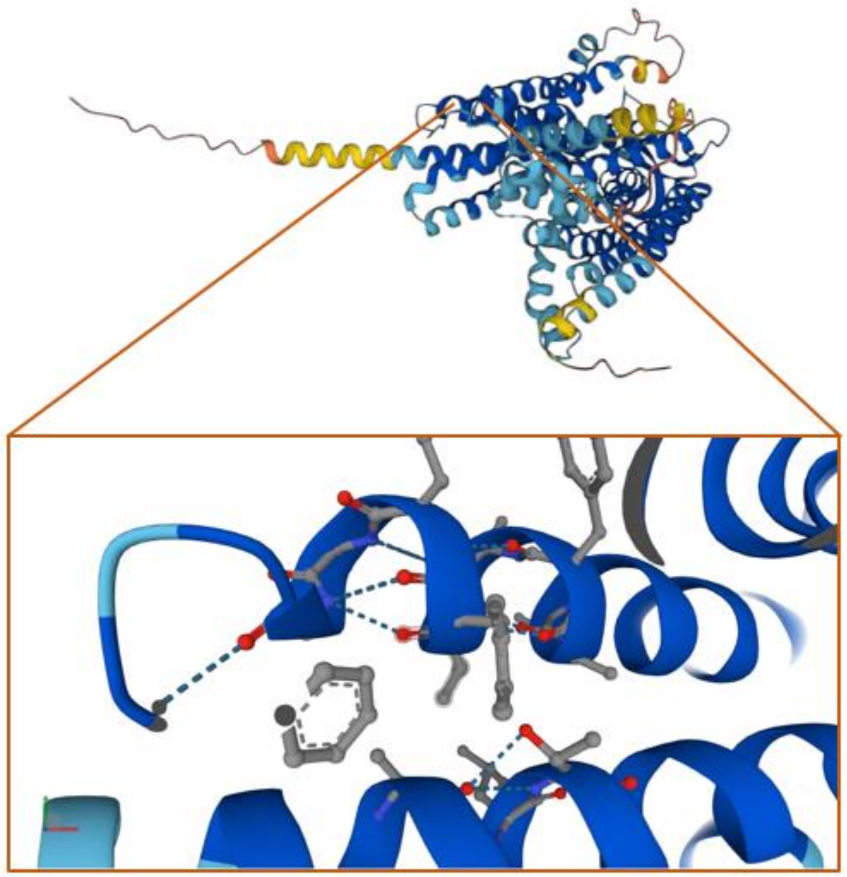

### Functional Dyspepsia GWAS - Garcia-Etxebarria et al.,- 2021

| chr | bp | rsID | Nearestgene | effect.allele | OR | P.logistic | MAF.gastro.all | MAF.non-gastro | MAF.diabetic | MAF.idiopathic | Mappedgenes |
|---|---|---|---|---|---|---|---|---|---|---|---|
| 1 | 204977799 | rs2595968 | NFASC | A | 0.74 | **0.0045** | 0.240 | 0.299 | 0.229 | 0.245 | NFASC, CNTN2, TMEM81, CDK18, MDM4, LRRN2 |
| 2 | 223923219 | rs6719560 | KCNE4 | T | 1.13 | 0.3645 | 0.129 | 0.115 | 0.138 | 0.124 | KCNE4, FARSB, PAX3, CCDC140, MOGAT1, SCG2, AP1S3 |
| 3 | 22296260 | rs144885331 | ZNF385D | C | 0.97 | 0.8946 | 0.033 | 0.034 | 0.024 | 0.037 | ZNF385D |
| 3 | 131682621 | rs9868674 | CPNE4 | T | 1.09 | 0.3625 | 0.436 | 0.415 | 0.467 | 0.421 | CPNE4, ASTE1, NEK11, MRPL3 |
| 3 | 172649336 | rs80062354 | SPATA16 | T | 1.07 | 0.5601 | 0.205 | 0.195 | 0.181 | 0.217 | SPATA16, NLGN1, NAALADL2 |
| 5 | 2703856 | rs961136 | IRX2 | T | 0.65 | 0.3066 | 0.011 | 0.017 | 0.010 | 0.012 | ADAMTS16 |
| 6 | 128345371 | rs17245411 | PTPRK | C | 1.07 | 0.7672 | 0.049 | 0.046 | 0.048 | 0.049 | PTPRK, THEMIS, LAMA2, TMEM244, L3MBTL3 |
| 7 | 8892730 | rs6463848 | | T | 1.00 | 0.9689 | 0.398 | 0.399 | 0.386 | 0.404 | NXPH1, NDUFA4, PHF14, THSD7A |
| 9 | 22206559 | rs9696092 | CDKN2B | C | 0.97 | 0.7137 | 0.425 | 0.433 | 0.395 | 0.439 | MTAP, RP11-145E5.5, C9orf53, CDKN2A, CDKN2B |
| 15 | 52868845 | rs184132620 | ARPP19 | C | 1.85 | 0.1182 | 0.017 | 0.009 | 0.024 | 0.014 | ARPP19, FAM214A, LYSMD2, SCG3 |
| 19 | 32610710 | rs17597505 | | T | 1.50 | **0.0383** | 0.066 | 0.045 | 0.076 | 0.061 | VSTM2B, POP4, C19orf12, CCNE1, URI1, ZNF536 |
| 20 | 61585706 | rs2093045 | SLC17A9 | C | 0.95 | 0.5891 | 0.364 | 0.376 | 0.314 | 0.388 | DIDO1, GID8, SLC17A9, COL9A3, TCLF5, BHLHE23 |
